# SARS-CoV-2 Serologic Assays in Control and Unknown Populations Demonstrate the Necessity of Virus Neutralization Testing

**DOI:** 10.1101/2020.08.18.20177196

**Authors:** Jennifer A. Rathe, Emily A. Hemann, Julie Eggenberger, Zhaoqi Li, Megan L. Knoll, Caleb Stokes, Tien-Ying Hsiang, Jason Netland, Kennidy Takehara, Marion Pepper, Michael Gale

**Author notes:** Corresponding author: Michael Gale, Jr. Building E (SLU 3.1) 750 Republican St. Room E360 | Box 35809 Seattle, WA 98109. These authors contributed equally to this study (Jennifer A. Rathe, Emily A. Hemann).

## Abstract

**Background:** To determine how serologic antibody testing outcome links with virus neutralization of SARS-CoV-2 to ascertain immune protection status, we evaluated a unique set of individuals for SARS-CoV-2 antibody detection and viral neutralization.

**Methods:** Herein, we compare several analytic platforms with 15 positive and 30 negative SARS-CoV-2 infected controls followed by viral neutralization assessment. We then applied these platforms in a clinically relevant population: 114 individuals with unknown histories of SARS-CoV-2 infection.

**Results:** In control populations, the best performing antibody detection assays were SARS-CoV-2 receptor binding domain (RBD) IgG (specificity 87%, sensitivity 100%, PPV 100%, NPV 93%), spike IgG3 (specificity 93%, sensitivity 97%, PPV 93%, NPV 97%), and nucleocapsid (NP) protein IgG (specificity 93%, sensitivity 97%, PPV 93%, NPV 97%). Neutralization of positive and negative control sera showed 100% agreement. 20 unknown individuals had detectable SARS-CoV-2 antibodies with 16 demonstrating virus neutralization. The antibody assays that best predicted virus neutralization were RBD IgG (misidentified 2), spike IgG3 (misidentified 1), and NP IgG (misidentified 2).

**Conclusion:** These data suggest that meaningful evaluation of antibody assay performance requires testing in an unknown population. Further, these results indicate coupling of virus neutralization analysis to a positive antibody test is required to categorize patients based on SARS-CoV-2 immune protection status following virus exposure or vaccine administration. One of the antibody detection platforms identified in this study followed by the pseudoneutralization or focus reduction assay would provide a practical testing strategy to assess for SARS-CoV-2 antibodies with optimal prediction of correlates to neutralizing immunity.

**Funding:** Supported by NIH grants AI148684, AI151698, AI145296, and UW funds to the Center for Innate Immunity and Immune Disease.

## Introduction

SARS-CoV-2 has infected millions of people globally but an accurate measure of the number of individuals with detectable neutralizing antibody titers within local and national populations remains unclear. Several clinically available serological assays to detect SARS-CoV-2 antibodies have been developed but their utility is hindered by limited sensitivity and specificity, which is worsened by the low prevalence of individuals within the population with SARS-CoV-2 antibodies(12, 13). Further, it is not clear how long neutralizing antibody titers are detectable after acute infection(16, 21). Therefore, interpretation of antibody results in the clinical setting is difficult such that individuals with waning antibodies, those without antibodies despite infection, and those with detectable antibodies but no neutralization activity cannot be identified using single tier serologic testing. Funding:

For most viral infections, threshold antibody titers have been established that correlate with protective immunity from symptomatic infection(3, 8, 9). Varicella zoster virus (VZV) vaccination studies have demonstrated the relationship between neutralizing antibodies and protection from symptomatic, acute VZV infection or reinfection(3, 8, 9, 17). Similar knowledge of how humoral immune responses to SARSCoV-2 infection relate to immune protection is developing rapidly, yet many questions remain. Recent studies of known SARS-CoV-2 infected individuals have demonstrated that detectable antibody levels are not always associated with viral neutralizing activity(12, 15, 24). However, there is emerging evidence that mildly symptomatic or asymptomatic individuals may have no or waning antibody titers over time(16, 21). However, reinfection with SARS-COV-2 following recovery from initial infection appears to be a rare event, as the few cases suggesting reinfection appear to be linked with the presence of residual viral RNA after recovery rather than a second infection with SARSCoV-2(14). Currently the best correlate of immunity is the presence of antibodies to the receptor binding domain of the SARS-CoV-2 spike protein that track with functional viral neutralization activity(11). The two-tiered testing approach to determine the presence of antibodies reactive to SARS-CoV-2 and subsequently, their ability to neutralize the virus, will provide much needed critical information to parse subjects into practical categories for future study and clinical interpretation. More importantly, coupling neutralization to clinical serologic testing will identify those individuals most likely to have virus neutralizing antibody titers suggesting immunity to SARS-CoV-2. Additionally, high-throughput interpretable serologic testing is critical for vaccine efficacy trials that are rapidly advancing and required to prevent future morbidity and mortality from SARS-CoV-2 infections.

The clinical testing market has been flooded with antibody detection platforms, displaying variable sensitivity and specificity(12, 13). The rapidity of production and testing of these platforms has been incredibly impressive but has exposed the high variability in performance of the different platforms(1, 12, 13, 18, 25). Further, the majority of testing to date has been completed on known positive and negative control subjects making interpretation of how each test will perform in random populations almost impossible. Given the relatively low prevalence of SARS-CoV-2 in most populations and less than ideal specificity of testing platforms, false positives are a significant problem(2). Platforms also differ in the viral antigens utilized for serologic testing. The most widely used viral antigens are the SARS-CoV-2 receptor binding domain (RBD) of the viral spike protein, the ectodomain of the spike protein, and nucleocapsid protein (NP). In conjunction, SARS-CoV-2 serologic tests generally assess the level of total IgG and/or IgM for antibody determination in an attempt to capture the time-dependent nature of antibody isotype reactivity; IgM is the first isotype produced, with IgG detectable at later time points post-infection and also serving as a marker of immune memory(10, 20). SARS-CoV-2 studies on antibody isotype time to detection have also been variable including IgG, IgM, IgG1, IgG2, IgG3, and IgA in positive control samples(1, 18, 23–25). While some studies have compared viral antigens and their reactivity with human antibody subtypes, it is not clear which combinations offer the most sensitive and specific serologic detection assay and most importantly, which combination best predicts viral neutralization in real world unknown subjects(15, 23, 24, 29).

In this study, our goal was to determine a serologic testing strategy least likely to identify false positives and most predictive of SARS-CoV-2 humoral immunity regardless of the time from acute infection. Therefore, we took a comprehensive approach comparing four viral antigens and five human antibody isotype ELISA platform combinations to determine which approach best predicts neutralizing antibody activity against SARS-CoV-2. We first assessed ELISA performance in positive and negative control SARS-CoV-2 infected groups. Subsequently, we tested the platforms performance in real-world unknown subjects. Lastly, we interpreted the serologic platforms’ performance in the context of detectable viral neutralization activity.

## Results

To evaluate the presence of SARS-CoV-2 specific antibodies and their neutralization capability, subjects were enrolled from the greater Seattle area for plasma collection and antibody testing. Positive controls consisted of 15 symptomatic RT-PCR SARS-CoV-2 positive individuals who were enrolled at least 21 days after the positive PCR test result. All of the positive controls had mild symptoms and none of the subjects were hospitalized, **Supplemental data file S1**. Thirty negative controls were obtained from a local blood bank as deidentified serum collected prior to December 1, 2019. Unknown samples consisted of two cohorts: G**roup A and Group B**. Group A includes 14 subjects with known exposure to confirmed SARS-CoV-2 infected, symptomatic individuals; the Group A subjects were never tested by PCR assay for determination of acute COVID-19 infection. Group B subjects were randomly recruited without knowledge of previous exposures to confirmed SARS-CoV-2 infected individuals. Subjects were recruited with posted flyers and websites, **Supplemental data file S1**. Serum samples were collected from March 26 to April 15, 2020.

Subjects were consented following protocols approved by the UW Institutional Review Board. Once enrolled in the study, information was collected from each subject including symptoms, zip code, sex, age, and results of any SARS-CoV-2 testing. Blood was collected, serum separated, heat inactivated, and stored at –80C.

ELISAs of serum from positive and negative controls were completed using 3 SARS-CoV-2 viral antigens: RBD, spike(26), and full SARS-CoV-2 UV-inactivated viral particles, **Figure 1A**. Serial dilutions of each sample generated area under the curve (AUC) values plotted with associated subject cohorts. Assessment of total IgG antibody levels for all 3 viral antigens displayed significant, distinct grouping of positive and negative controls. Antibody assessment by RBD and spike viral antigen ELISAs demonstrated the best separation between positive and negative controls. Contrastingly, ELISAs based on inactivated virus displayed the most overlap of reactivity by both positive and negative control cohorts. Given the comparatively improved identification of positive control samples by RBD and spike ELISA, unknowns were tested using these parameters with no further testing with UV-inactivated virus ELISA. Calculations for sensitivity, specificity, positive predictive value (PPV), negative predictive value (NPV), and associated 95% confidence intervals were determined based on the negative and positive control samples for ELISA platforms, **Table 1**. RBD IgG outperformed spike IgG in all measured parameters including specificity, sensitivity, PPV, and NPV. To assess the unknown samples (Group A and B)(23), a cut-off to identify samples as antibody-positive was calculated based on the mean plus 3 standard deviations of the negative control samples, designated as a dotted line on each graph. Unknowns assessed for IgG reactivity against RBD and spike identified 16 and 20 positive samples, respectively. Fifteen of the positive samples were identified by both platforms.

**Figure 1:**
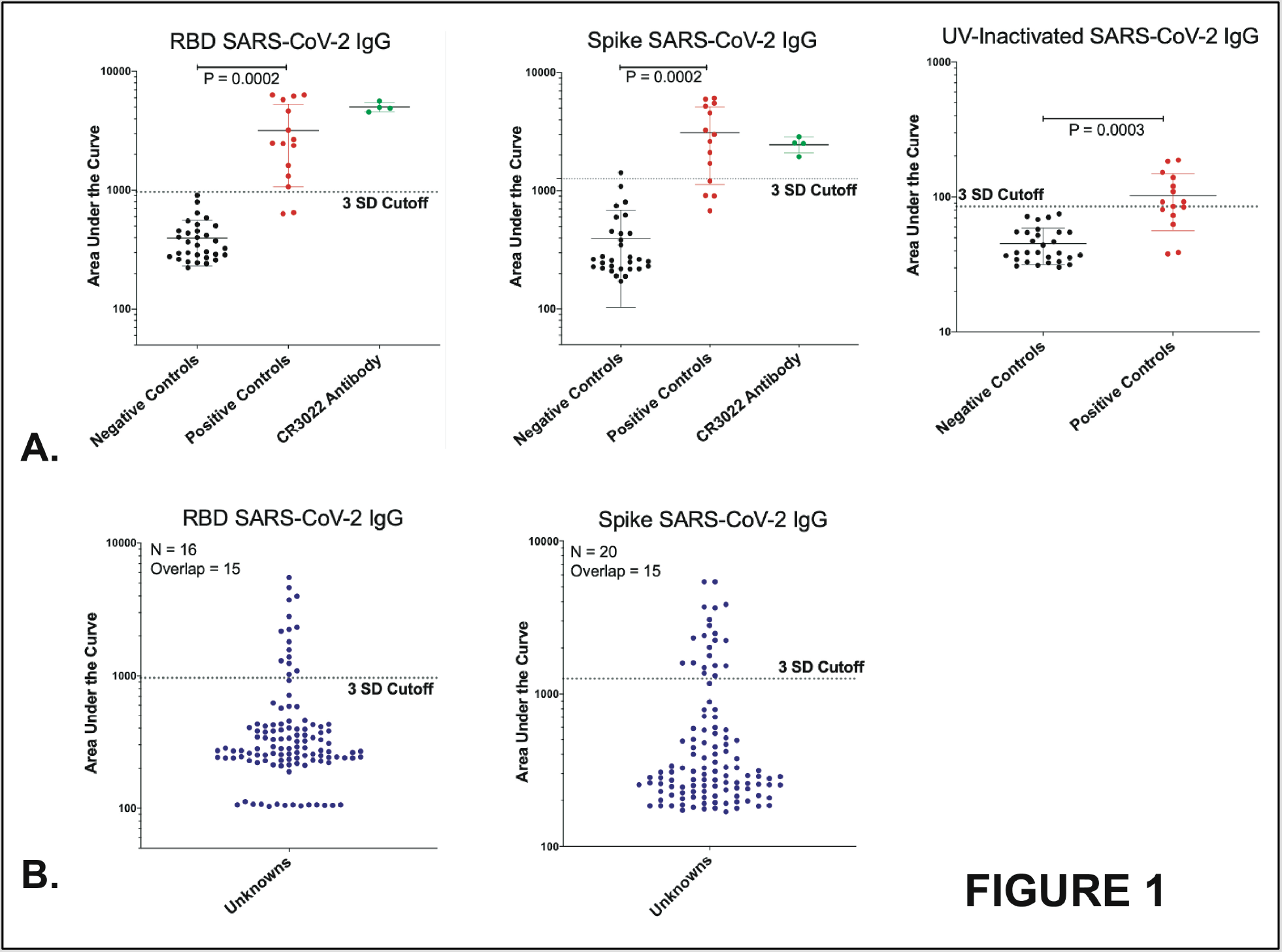
ELISA viral antigen comparisons. A.) Positive and negative control serum sample total IgG antibody recognition of SARS-CoV-2 RBD, spike protein, and UV-inactivated SARS-CoV-2. B.) Unknown serum sample total IgG RBD and spike assays. Mean +/− SD, Student’s t-tests for comparisons of mean of groups. Dotted lines represent cut-off of 3SD from the mean of the negative control samples to designate ‘positive’ antibody samples.

**Table 1:**
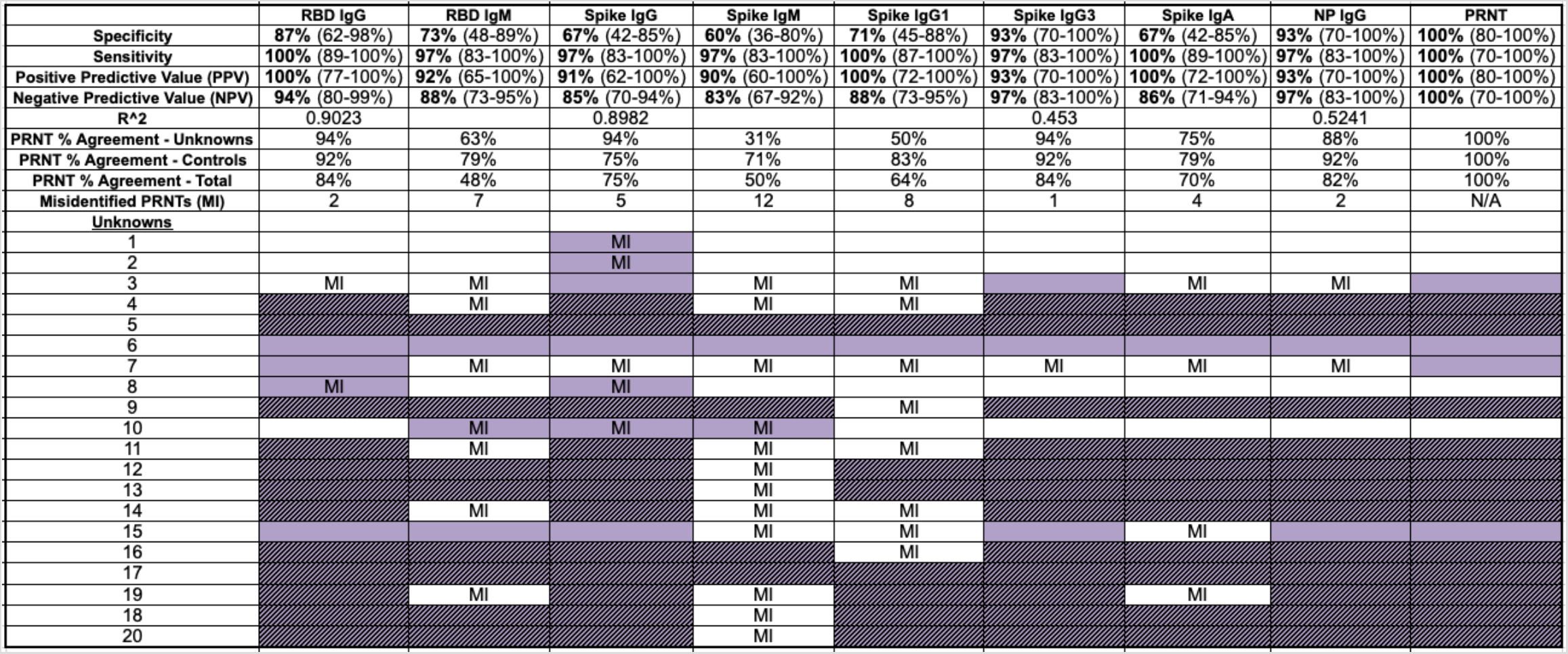
Summary of testing platform performance. Sensitivity, specificity, PPV, NPV, and associated 95% confidence intervals (CI) are based on positive and negative controls. 95% CIs are shown in parenthesis following the calculated values. PRNT performance and designation as ‘misidentified PRNTs’ based on the unknown samples meeting the neutralization designation in the setting ELISA platform recognition.

|  | RBD IgG | RBD IgM | Spike IgG | Spike IgM | Spike IgG1 | Spike IgG3 | Spike IgA | NP IgG | PRNT |
| --- | --- | --- | --- | --- | --- | --- | --- | --- | --- |
| **Specificity** | **87%** (62-98%) | **73%** (48-89%) | **67%** (42-85%) | **60%** (36-80%) | **71%** (45-88%) | **93%** (70-100%) | **67%** (42-85%) | **93%** (70-100%) | **100%** (80-100%) |
| **Sensitivity** | **100%** (89-100%) | **97%** (83-100%) | **97%** (83-100%) | **97%** (83-100%) | **100%** (87-100%) | **97%** (83-100%) | **100%** (89-100%) | **97%** (83-100%) | **100%** (70-100%) |
| **Positive Predictive Value (PPV)** | **100%** (77-100%) | **92%** (65-100%) | **91%** (62-100%) | **90%** (60-100%) | **100%** (72-100%) | **93%** (70-100%) | **100%** (72-100%) | **93%** (70-100%) | **100%** (80-100%) |
| **Negative Predictive Value (NPV)** | **94%** (80-99%) | **88%** (73-95%) | **85%** (70-94%) | **83%** (67-92%) | **88%** (73-95%) | **97%** (83-100%) | **86%** (71-94%) | **97%** (83-100%) | **100%** (70-100%) |
| **R^2** | 0.9023 |  | 0.8982 |  |  | 0.453 |  | 0.5241 |  |
| **PRNT % Agreement - Unknowns** | 94% | 63% | 94% | 31% | 50% | 94% | 75% | 88% | 100% |
| **PRNT % Agreement - Controls** | 92% | 79% | 75% | 71% | 83% | 92% | 79% | 92% | 100% |
| **PRNT % Agreement - Total** | 84% | 48% | 75% | 50% | 64% | 84% | 70% | 82% | 100% |
| **Misidentified PRNTs (MI)** | 2 | 7 | 5 | 12 | 8 | 1 | 4 | 2 | N/A |
| **Unknowns** |  |  |  |  |  |  |  |  |  |
| 1 |  |  | MI |  |  |  |  |  |  |
| 2 |  |  | MI |  |  |  |  |  |  |
| 3 | MI | MI |  | MI | MI |  | MI | MI |  |
| 4 |  | MI |  | MI | MI |  |  |  |  |
| 5 |  |  |  |  |  |  |  |  |  |
| 6 |  |  |  |  |  |  |  |  |  |
| 7 |  | MI | MI | MI | MI | MI | MI | MI |  |
| 8 | MI |  | MI |  |  |  |  |  |  |
| 9 |  |  |  |  | MI |  |  |  |  |
| 10 |  | MI | MI | MI |  |  |  |  |  |
| 11 |  | MI |  | MI | MI |  |  |  |  |
| 12 |  |  |  | MI |  |  |  |  |  |
| 13 |  |  |  | MI |  |  |  |  |  |
| 14 |  | MI |  | MI | MI |  |  |  |  |
| 15 |  |  |  | MI | MI |  | MI |  |  |
| 16 |  |  |  |  | MI |  |  |  |  |
| 17 |  |  |  |  |  |  |  |  |  |
| 19 |  | MI |  | MI |  |  | MI |  |  |
| 18 |  |  |  | MI |  |  |  |  |  |
| 20 |  |  |  | MI |  |  |  |  |  |

We next investigated the isotype diversity of the antibody response against SARS-CoV-2. For the positive and negative control samples collected in this study, IgM reactivity was decreased compared to IgG for both RBD and spike, **Figure 2A**. Comparison of assay performance measures between IgG and IgM assays for RBD and spike yielded relatively decreased sensitivity, specificity, PPV, and NPV for the IgM assays, **Table 1**. Analyses of the unknowns (Group A and B) with the IgM assays identified fewer positive samples compared to their IgG counterpart. However, all of the positives that were identified on the IgM assays were also identified on the IgG RBD and spike assays with one exception. There was one unknown sample identified as positive on the RBD IgM platform that was not identified on any other ELISA platform. To further investigate antibody isotype reactivity, all positive controls, negative controls, and unknowns identified as positive for IgG antibody titers against RBD or spike were additionally assessed for IgG1, IgG3, and IgA spike specific antibodies, **Figure 2B**. Of the antibody isotypes, IgG3 demonstrated the highest specificity and NPV while IgG1 and IgA had superior sensitivity and PPV. Spike IgG3 demonstrated a significantly better separation for the control samples: p = 0.0007 for IgG3 versus p = 0.02 and p = 0.002 for IgG1 and IgA, respectively. The spike IgG3 platform detected positive reactivity in 15 of the 20 unknown samples identified as positive by the IgG RBD and spike platforms.

**Figure 2:**
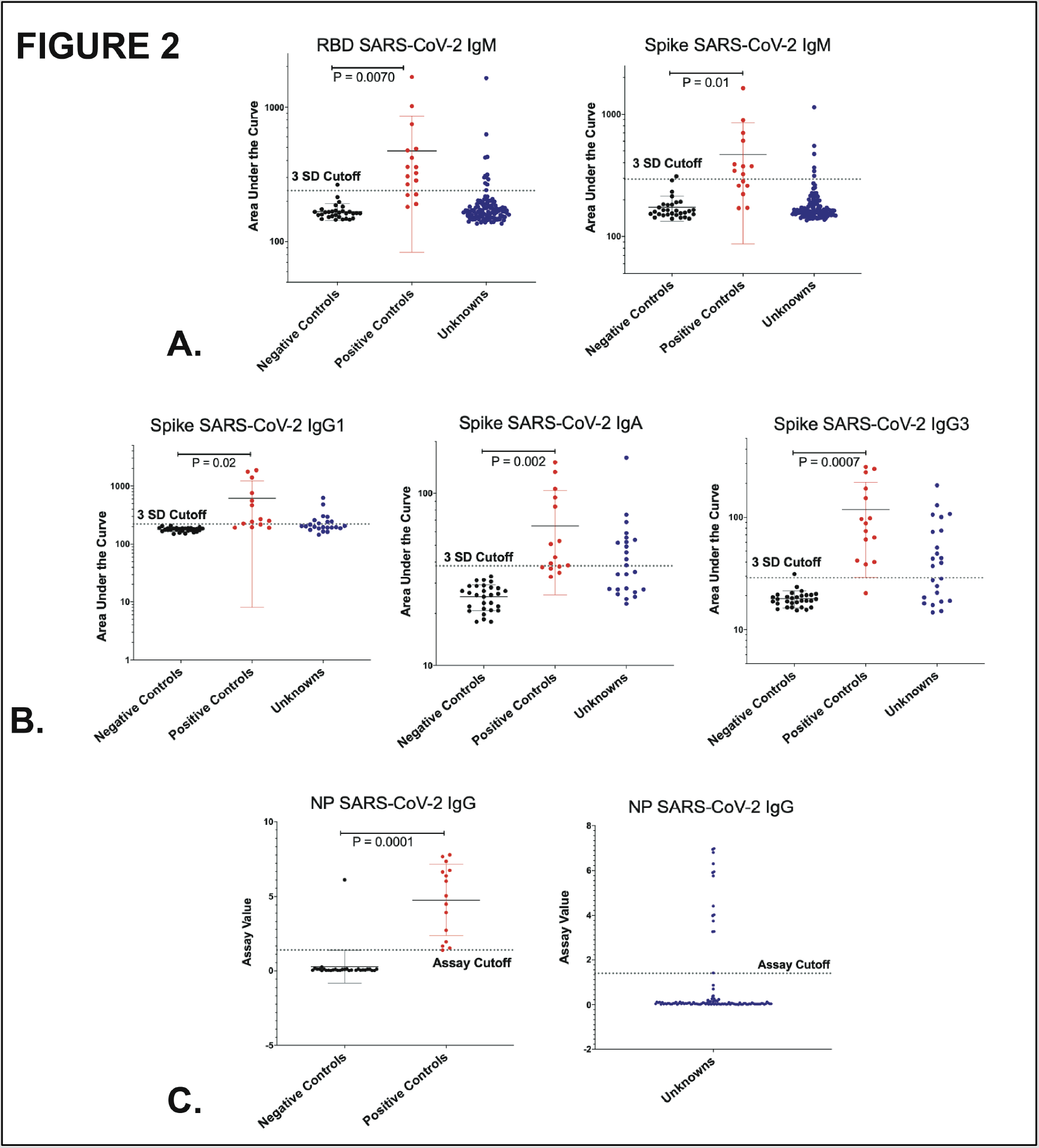
ELISA Antibody isotype comparisons. A.) IgM antibody recognition of SARSCoV-2 RBD and spike proteins of all serum samples. B.) IgG1, IgG3, and IgA antibody recognition of spike protein of all samples. Mean +/− SD, Student’s t-tests for comparisons of mean of groups. Dotted line represents cut-off of 3SD from the mean of the negative control samples to designate ‘positive’ antibody samples. C.) IgG SARSCoV-2 Abbott NP chemiluminescent microparticle immunoassay results, assay quantitative measurement output, > 1.39 = positive result.

We also tested our samples using an FDA emergency use authorized ELISA based on SARS-CoV-2 nucleoprotein (NP) platform (Abbot) to identify NP-specific IgG responses. The NP IgG platform resulted in similar significant separation of antibody-positive and negative subjects across our cohorts compared to serologic assessment against RBD IgG, spike IgG, and spike IgG3, p = 0.0001, **Figure 2C**. The NP IgG platform demonstrated similar sensitivity, specificity, PPV, and NPV compared to spike IgG3 and improved specificity compared to both RBD and spike IgG, **Table 1**. The NP IgG platform identified 14 unknowns as positive all of which were also identified across all of the following platforms: RBD IgG, spike IgG, and spike IgG3, **Table 1**.

Plaque reduction neutralization titer (PRNT) analysis against SARS-CoV2/ WA-1 isolate was used as to assess antibody function via virus neutralization activity, **Figure 3**. PRNT was performed on the following samples: all positive controls, 10 negative control samples (including five negatives identified as ‘positive’ on any platform), all RBD, spike, and NP IgG ‘positive’ unknowns, and 10 unknown samples near the cut-off but technically ‘negative’ for IgG RBD and spike. This selection of samples ensured neutralization would be assessed across the breadth of responses from each ELISA platform. For positive control subjects, we compared PRNT 80% reciprocal dilutions to ELISA AUC values for the platforms demonstrating the most significant separation of negative and positive control subjects, **Figure 3A**. RBD-specific IgG ELISA results show the highest correlation of the magnitude of ELISA antibody signal to the strength of neutralization followed by > spike IgG > NP > spike IgG3. We established a cutoff for identification of samples for positive neutralization: samples had to have detectable neutralization at both 50% (PRNT_50_) and 80% (PRNT_80_) with respective end point dilution set as 3SD greater than the mean value for negative control samples. **Figure 3B** demonstrates PRNT_80_ values in the left panel, with samples meeting the positive neutralization criteria in the right panel. All of the positive controls, 12 of the 14 Group A subjects, and 4 of the Group B subjects showed detectable neutralization above the cutoff criteria. Notably, none of the negative controls demonstrated neutralization.

**Figure 3:**
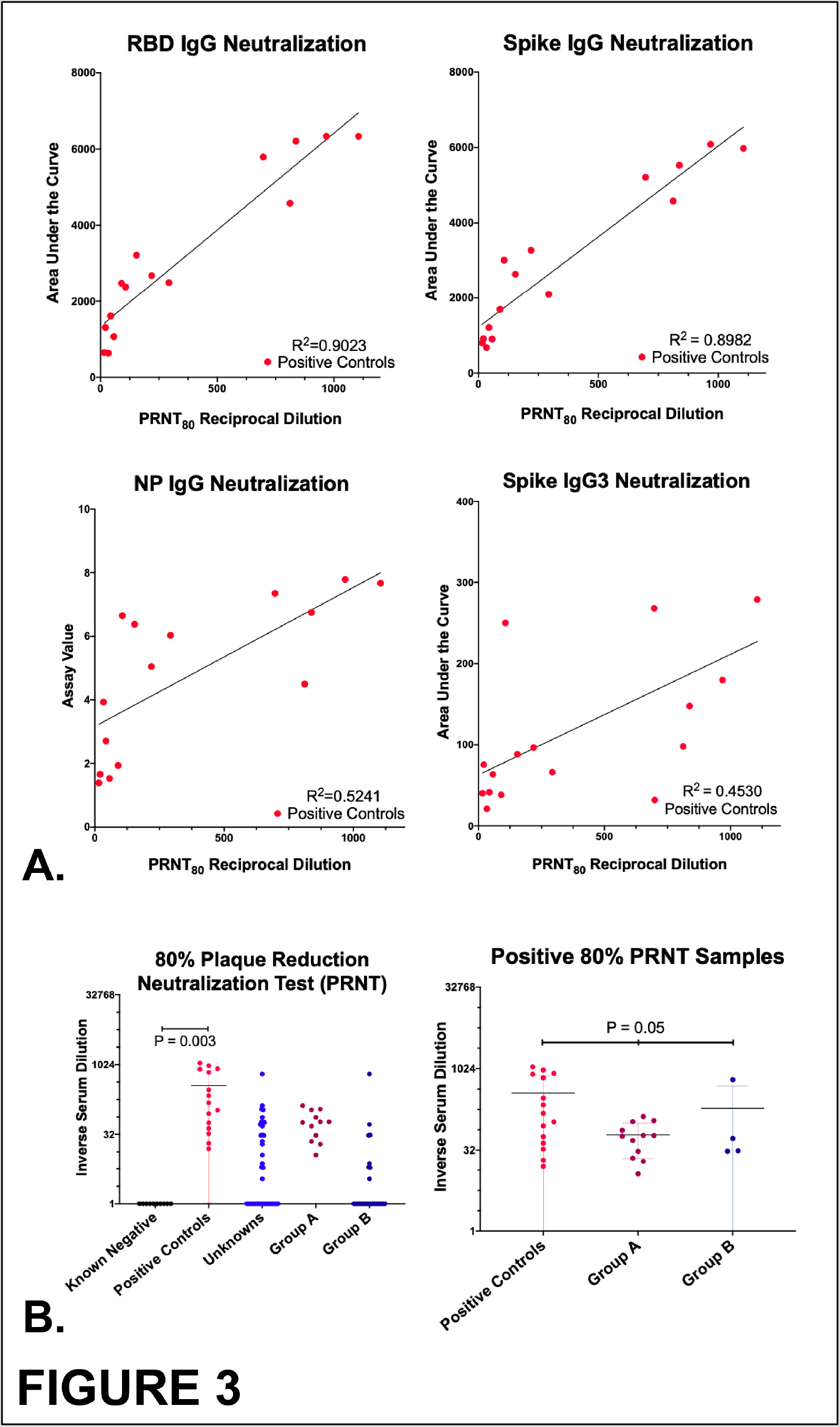
Plaque reduction neutralization test (PRNT) results. A.) Linear regression analyses correlating magnitude of neutralization PRNT_80_ to magnitude of ELISA serum: spike IgG, RBG IgG, NP IgG, and spike IgG3. B.) PRNT_80_ results for all samples and for those meeting neutralization criteria. Mean +/− SD, Student’s t-tests for comparisons of mean of two groups and ANOVA analyses for difference among means of three groups.

SARS-CoV-2 PRNT results provide key context to interpret ELISA results and provide functional evidence of viral neutralization. **Table 1** summarizes and compares measures of performance of the tested ELISA platforms on the control and unknown cohorts with correlation to the PRNT SARS-CoV-2 results. The first 9 rows show specific evaluations of the testing platforms assessed in this study. Sensitivity, specificity, PPV, NPV, and R^2^ measurements are based on the positive and negative control subjects’ antibody reactivity to each of the ELISA platform assays. Specificity and NPV are superior for spike IgG3 and NP IgG; therefore, these two platforms will identify fewer false positive samples compared to RBD and spike IgG. Sensitivity and PPV are best for RBD IgG, spike IgG1, and spike IgA platforms resulting in less false negative identifications. However, spike IgG1 and IgA suffer from much lower specificity and PPV compared to RBD IgG. Linking ELISA and the PRNT data, the R^2^ correlation between the magnitude of specific ELISA platform antibody detection and neutralization strength is highest for RBD and spike IgG. PRNT percent agreement was calculated for each platform as the number of subjects positive by specific ELISA platforms that were also positive for neutralization for controls samples and unknowns; RBD IgG and spike IgG3 had the highest percent agreement with PRNTs across all cohorts. Subject misidentifications were defined by discordant results between specific ELISA platforms and neutralization results that were not entirely accounted for by the PRNT percent agreement calculations. The spike IgG3 platform had the least number of misidentifications followed by NP IgG and RBD IgG. Sequential ELISA assay testing platforms have been proposed to increase sensitivity and specificity of SARS-CoV-2 antibody testing(23). **Table 2** compares sequential ELISA assays in terms of sensitivity, specificity, PRNT agreement and PRNT misidentifications. No two sequential ELISA assays outperformed RBD IgG, spike IgG3, or NP IgG. These results show that assessment of spike IgG3 antibodies provides the highest accuracy for revealing serologically positive individuals with neutralizing antibody with NP IgG and RBD IgG slightly inferior in their ability to predict neutralization in our sample set.

**Table 2:** Sequential ELISA assay analyses. Testing performance of two ELISA platforms requiring any positive result to be confirmed in the second platform to be considered a ‘true positive’. Solid color blocks = positive ELISA group B subject; solid color + crosshatch = positive ELISA, SARS-CoV-2 group A subject; PRNT% agreement = number of ELISA positives in agreement with PRNT positives.

|  | PRNT | Spike IgG3 | RBD IgG --> Spike IgG | RBD IgG --> Spike IgG3 | RBD IgG --> NP IgG | Spike IgG3 --> NP IgG |
| --- | --- | --- | --- | --- | --- | --- |
| **Specificity** | 100% | 93% | 67% | 87% | 87% | 87% |
| **Sensitivity** | 100% | 97% | 97% | 97% | 97% | 93% |
| **PRNT % Agreement - Unknowns** | 100% | 94% | 88% | 88% | 88% | 88% |
| **PRNT % Agreement - Controls** | 100% | 92% | 75% | 83% | 88% | 83% |
| **PRNT % Agreement - Total** | 100% | 84% | 73% | 77% | 80% | 77% |
| **Misidentified PRNTs (MI)** | N/A | 1 | 3 | 2 | 2 | 2 |
| **Unknowns** |  |  |  |  |  |  |
| 1 |  |  |  |  |  |  |
| 2 |  |  |  |  |  |  |
| 3 |  |  | MI | MI | MI | MI |
| 4 |  |  |  |  |  |  |
| 5 |  |  |  |  |  |  |
| 6 |  |  |  |  |  |  |
| 7 |  | MI | MI | MI | MI | MI |
| 8 |  |  | MI |  |  |  |
| 9 |  |  |  |  |  |  |
| 10 |  |  |  |  |  |  |
| 11 |  |  |  |  |  |  |
| 12 |  |  |  |  |  |  |
| 13 |  |  |  |  |  |  |
| 14 |  |  |  |  |  |  |
| 15 |  |  |  |  |  |  |
| 16 |  |  |  |  |  |  |
| 17 |  |  |  |  |  |  |
| 19 |  |  |  |  |  |  |
| 18 |  |  |  |  |  |  |
| 20 |  |  |  |  |  |  |

Using neutralization as the key designation of a positive unknown sample, we estimated the prevalence of individuals in the Seattle area with neutralizing antibodies against SARS-CoV-2 as of March-April 2020, **Figure 4**. Panel A is a map of the greater Seattle area with subjects designated by zip code. The majority of subjects were from within the city of Seattle. Panel B shows the number of positives identified from group A and B as well as the positives experiencing symptoms since the time of first documented human SARS-CoV-2 viral detection: January 21, 2020. Prevalence of neutralizing antibodies was estimated using a weighted logistic regression model adjusting for age and sex distributions according to King County census data, Panel C. The prevalence of individuals with SARS-CoV-2 antibodies at the time of this study is estimated at 3.5% with a 95% confidence interval range of 1.3 – 7.3%. Comparison of exposed and unexposed cohorts shows a highly significant difference in frequency of detection of SARS-CoV-2 antibodies, and reveals that if one has close interactions with a known infected individual in the absence of any measures to reduce transmission they are much more likely to develop detectable neutralizing antibodies.

**Figure 4:**
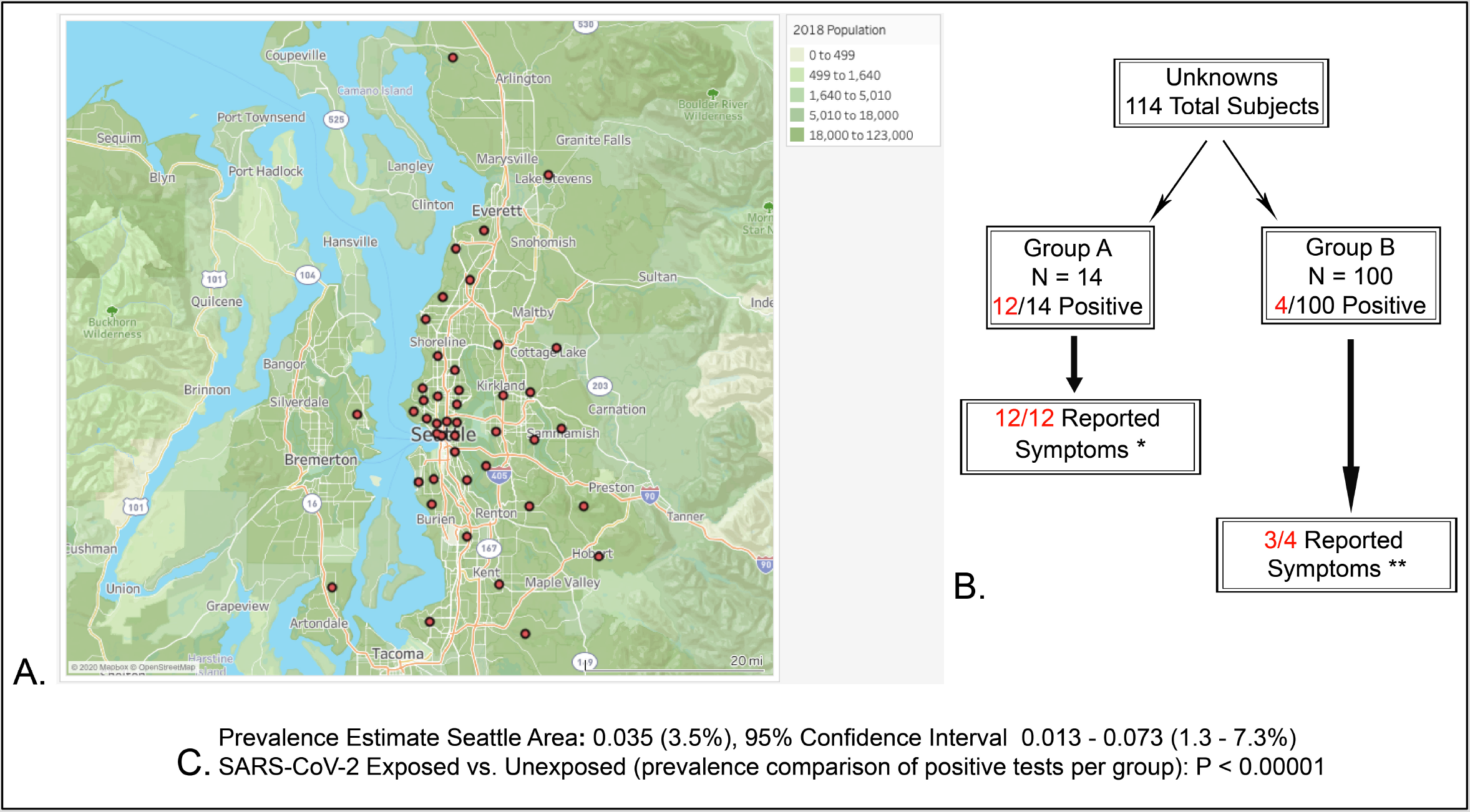
Prevalence estimate and subject details. A.) A map of Seattle with circles denoting the zip codes of the subjects involved in the study. Underlying map colors represent the population density for the areas shown. B.) Boxes demonstrate the number of subjects, exposure status, and reported symptoms for positive subjects. Group A and B unknown subjects have never been diagnosed with SARS-CoV-2 infection. Group A subjects had close contact with a known infected SARS-CoV-2 individual. Group B subjects had no known exposure to SARS-CoV-2 infected individuals. C.) The estimated prevalence of neutralizing SARS-CoV-2 antibodies in the greater Seattle area with associated 95% CI. The p value calculated from a comparison of the exposed versus unexposed groups proportion of positive test results in our study. * Subjects reported symptoms within 5 days of exposure to SARS-CoV-2 positive individuals. ** Subjects reported symptoms over the possible exposure window in the Seattle area (Jan 21, 2020 – April 15, 2020).

## Discussion

Our study shows that a two-tiered testing strategy of ELISA followed by PRNT of positive ELISA samples is the most accurate and efficient way to assess humoral immunity to SARS-CoV-2. We completed comprehensive analyses of multiple ELISA SARS-CoV-2 platforms coupled with the gold standard of viral neutralization testing to determine a testing strategy that was least likely to identify false positives and most predictive of SARS-CoV-2 neutralization regardless of the time from acute infection. Relying on results from a single analysis of testing performance (i.e. a single row in **Table 1**) does not provide a comprehensive assessment of the accuracy of the ELISA platforms. Furthermore, we found that having paired neutralization studies was key to interpreting results of the ELISAs, particularly when assessing an unknown population.

Three ELISA platforms rise to the top of our list based on our desire for high specificity and NPV, high PRNT agreement, and low misidentification of subjects with incongruent ELISA and neutralization results: RBD IgG, spike IgG3, and NP IgG. Spike IgG3 surpasses RBD IgG because of its superior specificity and therefore a lower likelihood of false positives as well as its superior prediction of neutralization due to the lower number of misidentifications compared to RBD IgG. Of note, the R^2^ for RBD IgG was relatively higher compared to the lower correlation for spike IgG3. However, this is challenging to interpret given the finding in other studies with little correlation with RBD IgG to neutralization (R^2^ ranging from 0.5 – 0.7) when assessed in larger positive control groups(24, 28). Our positive controls were all collected around the same time from infection onset and may explain our observed high correlation. Based on our results, we conclude that R^2^ calculations alone are not very informative to assess ELISA performance for SARS-CoV-2. In assessing unknown populations with our platforms, we found that despite the higher correlation with PRNT, RBD IgG performed somewhat less well at predicting neutralization on a patient-by-patient basis as compared to spike IgG3, which had a comparatively low R^2^. With the variable reported values of R^2^ for RBD and spike platforms, this does not appear to be a dependable measurement of performance for SARS-CoV-2 ELISA platforms to predict neutralization.

Across the ELISA platforms we investigated, our data supports the use of spike-reactive IgG3 as the best initial screening test to predict neutralization but with the important caveat of our limited sample size. Many of the parameters of test performance were similar for RBD IgG, spike IgG3, and NP IgG. However, spike IgG3 edged out both NP and RBD IgG with the ability to predict neutralization (% PRNT agreement and misidentification columns in **Table 1**). Expanded studies comparing these three platforms with larger sample sizes would very beneficial to assess if spike IgG3 continues to outperform the other two platforms. RBD and NP IgG platforms already have FDA emergency approval, with the NP IgG platform having the highest specificity and most comprehensive validation(4). Spike IgG3 platforms have not been developed but represent a highly promising platform for development given the knowledge that IgG3 isotypes are known as the most effective at viral neutralization compared to other IgG subtypes(10, 20). The NP IgG assay is likely to be limited in its ability to predict neutralization due to its localization within the virion: the NP protein is hidden beneath the viral envelope and it is therefore unlikely to be an effective target for neutralizing antibodies, unlike the spike proteins(27). However coupling the existing NP IgG Abott ELISA detection assay with neutralization would be a rapid and practical way to address this possible issue.

Classic PRNTs are expensive and time-consuming mostly due to laboratory biosafety requirements. However, PRNTs are currently the most accurate *in vitro* assay to functionally assess for the ability of human antibodies to neutralize live virus. Recent work has demonstrated high correlation between SARS-CoV-2 PRNT assay and focus reduction (FRNT) assay or neutralization utilizing a fluorescent SARS-CoV-2, which allows for a high-throughput sample analysis of neutralizing antibodies(19, 24). In addition, the application of pseudovirus neutralization assays can reduce the barrier to testing that requires access to biosafety level three facilities(6). In this study, we show how neutralizing assays serve as a check on the accuracy of antibody screening tests prone to false positives and negatives. Rapid, significant contributions by scientists worldwide have produced serologic SARS-CoV-2 data that has been consistent in one very important way: variability. Our study confirms high variability in serologic assay platform performance and provides further data that no single serologic assay provides perfect prediction for viral neutralizing ability. Two-tiered testing would allow parsing of subjects into groups important for further study (1) those with positive ELISA antibody detection and confirmed neutralization and (2) those with positive ELISA antibody detection but no evidence of neutralization. Group 1 would allow tracking of subjects with known neutralization titers for evidence of reinfection vs. protective immunity. Group 2 would allow study for increased identification of true false positives vs. individuals that do not develop functional and/or lasting neutralizing antibodies. Therefore, a two-tiered testing strategy in which high-throughput neutralization studies are coupled to serologic assays will provide fully interpretable data to identify subjects that have truly been infected with SARS-CoV-2, those with protective immunity, and to assess vaccine efficacy.

Lastly, we wanted to estimate the prevalence of individuals with detectable SARS-CoV-2 neutralizing antibodies in the Seattle area. Our samples were drawn in late March to early April. Our estimate of 3.5% prevalence in the Seattle area is similar to other published studies on the West Coast(22). Our sample size is quite small and therefore, unavoidably biased. The CDC reported a prevalence of 1.13% in samples obtained from the same time frame but their study also suffers from sampling bias(5). While samples were collected from a much larger catchment area compared to our study, blood bank donation samples were used rather than random enrollment(5). We propose that the 95% confidence interval range found in this study is perhaps a better representative of the true prevalence in our community (1.3 – 7.3%) rather than one specific value.

To our knowledge this is the first study to comprehensively assess serologic assay performance in an unknown cohort in addition to negative and positive controls in order to determine a practical and accurate method to determine the presence of SARS-CoV-2 neutralizing antibodies. By coupling ELISA with virus neutralization assessment, we were able to determine the accuracy of testing based on the key functional outcome of viral neutralization. Pseudoneutralization or FRNT assays provide a comparable test of neutralization to PRNTs and could be rapidly developed with a new spike IgG3 or existing NP IgG ELISA platform for two-tiered testing(6, 19, 24). Our study provides compelling evidence that a two-tiered neutralization-dependent testing scheme is required to correctly identify and categorize individuals with and without detectable neutralization to study the range of antibody responses to infection, the correlate to protective immunity, and vaccine efficacy.

## Methods

### Sample collection

Venipuncture collected 6–10 mls of blood in EDTA blood collection tubes. The tubes were spun at 1000 rcf for 10 minutes. Serum was collected and stored at –80C.

### ELISA

Elisa assays were performed as previously described (23). Briefly, high-binding plates (ThermoScientific) were coated with SARS-CoV-2 RBD (23), SARS-CoV-2 spike(26), or UV-inactivated SARS-CoV-2 (WA1, BEI resources) and incubated overnight at 4° C. Plates were blocked in PBST + 3% milk for 1hr at RT. Three-fold serial dilutions of heat-inactivated plasma were added to plates in biological duplicates. Consistent historical negative samples and the positive control spike-binding antibody CR3022 (Abcam, ab273073) were included on plates with total IgG antibody binding for specific viral antigen and antibody combinations. UV-inactivated SARS-CoV-2 did not demonstrate binding. Following a 2hr incubation and washes, the following anti-human secondary antibodies conjugated to HRP were diluted 1:3000 and added to plates: IgG (Thermofisher 31410), IgG1 (Southern Biotech 9054), IgG3 (Southern Biotech 9210), IgM (Sigma A6907), IgA (Sigma A0295). Following a 1hr incubation and washes, SigmaFast OPD was added to plates. Ten minutes later, 2M H_2_SO_4_ was added to all wells to stop the reaction and plates were immediately read at an absorbance of 490nm (BioTek Epoch). OD values for each sample dilution were plotted and the area under the curve (AUC) was calculated using Prism.

### PRNT Assay

PRNT assays were performed as previously described (7). Briefly, fourfold serial dilutions of heat inactivated plasma was mixed 1:1 with 600 PFU/ml SARSCoV-2 WA-1 (BEI resources) in DPBS (Fisher Scientific) + 0.3% cold water fish skin gelatin (Sigma G7041) and incubated for 30 min at 37 degrees. The virus plasma mixture was then added in duplicate, along with virus only and mock controls, to Vero cells (ATCC) in a 12-well plate and incubated for 1hr at 37 degrees. Following adsorption, plates were washed once with DPBS and overlayed with a 1:1 mixture of 2.4% Avicel RC-591 (FMC) + 2 x MEM (ThermoFisher) supplemented with 4% heat-inactivated FBS and Penicillin/Streptomycin (Fisher Scientific). Plates were then incubated for 2 days at 37 degrees. Following incubation, overlay was removed and plates were washed once with DPBS and fixed in 10% formaldehyde (Sigma-Aldrich) in DPBS for 30 minutes at room temp. Plates were washed again with DPBS and stained with 1% crystal violet (Sigma-Aldrich) in 20% EtOH (Fisher Scientific). Plaques were enumerated and percent neutralization was calculated as 100 minus the number of plaques in serum + virus dilution wells divided by the number of plaques in virus only control wells times 100. PRNT50 and PRNT80 values are shown as inverse serum dilution and were determined by calculating the 50% and 80% sigmoidal interpolation of the percent neutralization of the samples in Prism. R^2^ values were determined using a nonlinear regression fit in Prism.

### University of Washington NP Assay

Abbott SARS-CoV-2 IgG Testing Serum samples were run on the Abbott Architect instrument using the Abbott SARS-CoV-2 IgG assay after FDA notification following manufacturer’s instructions. The assay is a chemiluminescent microparticle immunoassay for qualitative detection of IgG in human serum or plasma against the SARS-CoV-2 nucleoprotein. The Architect requires a minimum of 100µL of serum or plasma. Qualitative results and index values reported by the instrument were used in analysis. Values > / = 1.4 is considered a positive result.

### Statistics

#### Prevalence Estimate

We first compare the age and sex distributions of the 100 unexposed unknown subjects and the age and sex distributions of the greater Seattle area from the U.S. Census estimates from 2014–2018. By using the Pearson’s chi-square test for sex and Kolmogorov-Smirnov (KS) test for age, we found significant differences for sex and age distribution between the unexposed subjects and the greater Seattle area. Looking at the histogram, there are more old people and less young people in the subjects comparing to people in Seattle area.

Therefore, we adjust this oversampling and under-sampling by constructing weights for age brackets and sex designations. We construct weights by taking the ratio of the proportion of the Seattle area versus the sample proportion. A logistic regression using the weight adjustments for sex and age was then conducted to calculate a prevalence estimate and the associated 95% confidence interval. As a result, the model gives a point estimate for the prevalence of 3.5% with a 95% confidence interval from 1.3% to 7.3%.

#### Comparison testing of Group A and Group B

We use the Fisher’s exact test to compare the rate of positive neutralizing antibodies against SARS-CoV-2 between the exposed and unexposed groups. The p-value of the Fisher exact test is less than 10^−5^, which is statistically significant. The estimated difference of the rate between two groups is –0.89 with a 95% confidence interval of (−0.97, –0.66), while it should be noted that the rates are between 0 and 1. Therefore, we can see that the rates between the two exposure groups are very different. <u>Further specifics are included in supplemental data file 2, S2</u>.

#### Sensitivity, specificity, PPV, NPV, and associated confidence intervals

The values for sensitivity, specificity, PPV, and NPV for the ELISA platforms and PRNT were calculated using the positive and negative controls as reference samples. Prism software contingency tables were used to calculate the specific values and 95% confidence intervals. Fisher’s exact test computed p-values and significance. The 95% confidence intervals were calculated using the hybrid Wilson/Brown method.

### Human participants

The institutional review board at the University of Washington reviewed and approved our human subjects study prior to enrollment of any subjects. Subjects were provided with information about the study, risks associated, and how their privacy would be protected. To enroll in the study each subject was required to provide verbal understanding and written consent. The IRB approved all recruitment and consent forms prior to the beginning of the study.

## Data Availability

All data that is not included in the manuscript and supplemental material is available upon request to interested parties. Data will not be released if it violates the privacy of our subjects as required by our IRB.

## Author Contributions

JAR – designed research study, acted as study coordinator, conducted experiments, acquired data, analyzed data, and wrote the manuscript

EAH – designed research study, conducted experiments, acquired data, analyzed data, and edited the manuscript

Co-first authors: JAR and EAH were both required for completion of this study and shared management duties throughout the project. JAR initially proposed the study and managed the clinical portion of the study. EAH developed and managed the laboratory experimental portions of the study.

JE and ML – conducted experiments, acquired data, analyzed data, and edited the manuscript

ZL – designed the research study, analyzed data, and edited the manuscript

TYH – conducted experiments, provided reagents, and edited the manuscript

CS and KT – coordinated sample collection from subjects, acquired data, and edited the manuscript

JN and MO – designed research study, analyzed data, provided reagents, and edited the manuscript

MG – designed research study, provided funding for the study, and edited the manuscript

## Acknowledgements

Supported by NIH grants AI148684, AI151698, AI145296, and UW funds to the Center for Innate Immunity and Immune Disease. We thank Keith Jerome and Gregory Pepper for SARS-CoV-2 NP ELISA. We thank Neil King and his lab members for providing soluble trimeric SARS-CoV-3 spike protein used in our ELISA assays. We thank Lauren Rodda for her input on experimental design and analyses.

## References

1. Beavis KG, Matushek SM, Abeleda APF, Bethel C, Hunt C, Gillen S, et al. Evaluation of the EUROIMMUN Anti-SARS-CoV-2 ELISA Assay for detection of IgA and IgG antibodies. J Clin Virol. 2020 May;129:104468.

2. Bentley TG, Catanzaro A, Ganiats TG. Implications of the impact of prevalence on test thresholds and outcomes: lessons from tuberculosis. BMC Res Notes. 2012 Oct;5:563.

3. Breuer J, Schmid DS, Gershon AA. Use and limitations of varicella-zoster virus-specific serological testing to evaluate breakthrough disease in vaccinees and to screen for susceptibility to varicella. J Infect Dis. 2008 Mar;197 Suppl 2:S147–51.

4. Bryan A, Pepper G, Wener MH, Fink SL, Morishima C, Chaudhary A, et al. Performance Characteristics of the Abbott Architect SARS-CoV-2 IgG Assay and Seroprevalence in Boise, Idaho. J Clin Microbiol. 2020 May.

5. CDC. Commercial Laboratory Seroprevalence Study. CDC2020 [cited 2020].

6. Crawford KHD, Eguia R, Dingens AS, Loes AN, Malone KD, Wolf CR, et al. Protocol and Reagents for Pseudotyping Lentiviral Particles with SARS-CoV-2 Spike Protein for Neutralization Assays. Viruses. 2020 05;12(5).

7. Erasmus JH, Khandhar AP, Walls AC, Hemann EA, O’Connor MA, Murapa P, et al. Single-dose replicating RNA vaccine induces neutralizing antibodies against SARS-CoV-2 in nonhuman primates. bioRxiv. 2020 May.

8. Gershon AA, Larussa P, Steinberg S. Detection of antibodies to varicella-zoster virus using a latex agglutination assay. Clin Diagn Virol. 1994 Aug;2(4–5):271–7.

9. Hammond O, Wang Y, Green T, Antonello J, Kuhn R, Motley C, et al. The optimization and validation of the glycoprotein ELISA assay for quantitative varicella-zoster virus (VZV) antibody detection. J Med Virol. 2006 Dec;78(12):1679–87.

10. Irani V, Guy AJ, Andrew D, Beeson JG, Ramsland PA, Richards JS. Molecular properties of human IgG subclasses and their implications for designing therapeutic monoclonal antibodies against infectious diseases. Mol Immunol. 2015 Oct;67(2 Pt A):171–82.

11. Ju B, Zhang Q, Ge J, Wang R, Sun J, Ge X, et al. Human neutralizing antibodies elicited by SARSCoV-2 infection. Nature. 2020 May.

12. Kohmer N, Westhaus S, Rühl C, Ciesek S, Rabenau HF. Brief clinical evaluation of six high-throughput SARS-CoV-2 IgG antibody assays. J Clin Virol. 2020 Jun;129:104480.

13. Kohmer N, Westhaus S, Rühl C, Ciesek S, Rabenau HF. Clinical performance of different SARSCoV-2 IgG antibody tests. J Med Virol. 2020 Jun.

14. Lan L, Xu D, Ye G, Xia C, Wang S, Li Y, et al. Positive RT-PCR Test Results in Patients Recovered From COVID-19. JAMA. 2020 Feb.

15. Long QX, Liu BZ, Deng HJ, Wu GC, Deng K, Chen YK, et al. Antibody responses to SARS-CoV-2 in patients with COVID-19. Nat Med. 2020 Apr.

16. Long QX, Tang XJ, Shi QL, Li Q, Deng HJ, Yuan J, et al. Clinical and immunological assessment of asymptomatic SARS-CoV-2 infections. Nat Med. 2020 Jun.

17. Ludwig B, Kraus FB, Allwinn R, Keim S, Doerr HW, Buxbaum S. Loss of varicella zoster virus antibodies despite detectable cell mediated immunity after vaccination. Infection. 2006 Aug;34(4):222–6.

18. Montesinos I, Gruson D, Kabamba B, Dahma H, Van den Wijngaert S, Reza S, et al. Evaluation of two automated and three rapid lateral flow immunoassays for the detection of anti-SARS-CoV-2 antibodies. J Clin Virol. 2020 May;128:104413.

19. Muruato AE, Fontes-Garfias CR, Ren P, Garcia-Blanco MA, Menachery VD, Xie X, et al. A high-throughput neutralizing antibody assay for COVID-19 diagnosis and vaccine evaluation. bioRxiv. 2020 May.

20. Papadea C, Check IJ. Human immunoglobulin G and immunoglobulin G subclasses: biochemical, genetic, and clinical aspects. Crit Rev Clin Lab Sci. 1989;27(1):27–58.

21. Seow J, Graham C, Merrick B, Acors S, Steel KJA, Hemmings O, et al. Longitudinal evaluation and decline of antibody responses in SARS-CoV-2 infection. medRxiv. 2020:2020.07.09.20148429.

22. Sood N, Simon P, Ebner P, Eichner D, Reynolds J, Bendavid E, et al. Seroprevalence of SARS-CoV-2-Specific Antibodies Among Adults in Los Angeles County, California, on April 10–11, 2020. JAMA. 2020 May.

23. Stadlbauer D, Amanat F, Chromikova V, Jiang K, Strohmeier S, Arunkumar GA, et al. SARS-CoV-2 Seroconversion in Humans: A Detailed Protocol for a Serological Assay, Antigen Production, and Test Setup. Curr Protoc Microbiol. 2020 06;57(1):e100.

24. Suthar MS, Zimmerman M, Kauffman R, Mantus G, Linderman S, Vanderheiden A, et al. Rapid generation of neutralizing antibody responses in COVID-19 patients. medRxiv. 2020 May.

25. Van Elslande J, Houben E, Depypere M, Brackenier A, Desmet S, André E, et al. Diagnostic performance of 7 rapid IgG/IgM antibody tests and the Euroimmun IgA/IgG ELISA in COVID-19 patients. Clin Microbiol Infect. 2020 May.

26. Walls AC, Park YJ, Tortorici MA, Wall A, McGuire AT, Veesler D. Structure, Function, and Antigenicity of the SARS-CoV-2 Spike Glycoprotein. Cell. 2020 04;181(2):281–92.e6.

27. Weiss SR, Leibowitz JL. Coronavirus pathogenesis. Adv Virus Res. 2011;81:85–164.

28. Wu F, Wang A, Liu M, Wang Q, Chen J, Xia S, et al. Neutralizing antibody responses to SARS-CoV-2 in a COVID-19 recovered patient cohort and their implications. medRxiv. 2020:2020.03.30.20047365.

29. Xiang F, Wang X, He X, Peng Z, Yang B, Zhang J, et al. Antibody Detection and Dynamic Characteristics in Patients with COVID-19. Clin Infect Dis. 2020 Apr.

